# Disparities in Case Frequency and Mortality of Coronavirus Disease 2019 (COVID-19) Among Various States in the United States

**DOI:** 10.1101/2020.07.28.20163931

**Authors:** Rohit S. Loomba, Gaurav Aggarwal, Saurabh Aggarwal, Saul Flores, Enrique G. Villarreal, Juan S. Farias, Carl J. Lavie

**Affiliations:** Advocate Children’s Hospital, Chicago, IL, USA; Chicago Medical School/ Rosalind Franklin University of Medicine and Science, Chicago, IL, USA; Jersey City Medical Center, Jersey City, NJ, USA; UnityPoint Health, Des Moines, IA, USA; Texas Children’s Hospital, Houston, TX, USA; Baylor College of Medicine, Houston, TX, USA; Tecnologico de Monterrey, Escuela de Medicina y Ciencias de la Salud, Monterrey, Nuevo Leon, Mexico; Ochsner Medical Center, New Orleans, LA, USA

**Keywords:** COVID-19, coronavirus, climate, risk factors, epidemiology

## Abstract

**Objective:** To utilize publicly reported, state-level data to identify factors associated with the frequency of cases, tests, and mortality in the US.

**Materials & Methods:** Retrospective study using publicly reported data collected included the number of COVID-19 cases, tests, and mortality from March 14^th^ through April 30^th^, 2020. Publicly available state-level data was collected which included: demographics comorbidities, state characteristics and environmental factors. Univariate and multivariate regression analyses were performed to identify the significantly associated factors with percent mortality, case and testing frequency. All analyses were state-level analyses and not patient-level analyses.

**Results:** A total of 1,090,500 COVID-19 cases were reported during the study period. The calculated case and testing frequency were 3,332 and 19,193 per 1,000,000 patients. There were 63,642 deaths during this period which resulted in a mortality of 5.8%. Factors including to but not limited to population density (beta coefficient 7.5, p< 0.01), transportation volume (beta coefficient 0.1, p< 0.01), tourism index (beta coefficient -0.1, p=0.02) and older age (beta coefficient 0.2, p=0.01) are associated with case frequency and percent mortality.

**Conclusions:** There were wide variations in testing and case frequencies of COVID-19 among different states in the US. States with higher population density had a higher case and testing rate. States with larger population of elderly and higher tourism had a higher mortality.

**Key Messages:** There were wide variations in testing and case frequencies of COVID-19 among different states in the US.

States with higher population density had a higher case and testing rate.

States with larger population of elderly and higher tourism had a higher mortality.

## Introduction

The coronavirus disease 2019 (COVID-19) has spread worldwide after its onset in Wuhan, China, reaching over three million cases^1^. In the United States (US), over seven million tests have been performed with over one million tests resulting positive. This disease has resulted in over 70,000 deaths in the US and a case fatality rate at approximately 6.2%. Recent reports have shown differences in case and fatality rates between nearby localities and thought to be likely due to differences in demographic and socioeconomic factors^2^. Furthermore, differences in climate, access to healthcare and adherence to social distancing have also been hypothesized to affect these rates^3–5^. However, factors mediating the differences in case frequency and testing frequency between the states in the US have not been formally assessed. The aim of this study is to use publicly available, state-level data to determine the factors associated with COVID-19 percent mortality and case and testing frequency.

## Materials and Methods

This study utilized publicly available, deidentified, state-level data and so no institutional review board approval was required or sought. The study was performed in accordance with Declaration of Helsinki.

### Variable Identification and Data Collection

Absolute counts for COVID-19 cases, tests, and mortality were obtained from Worldometer^6^. Data were collected from March 13^th^, 2020 through April 30^th^, 2020. These dates were selected as March 13^th^ was the first day when these data were publicly reported, and April 30th was the last full day prior to data collection. The number of cases and tests were then normalized to the specific state’s population to develop a frequency per 1,000,000 population. Any reference in this manuscript to “case frequency” or “testing frequency” refers to the normalized values in this manner, unless explicitly stated otherwise.

Next, data were collected at a state-wide level to help characterize the population, environment, and infrastructure in the state. The data sources are listed in Supplementary file 1. The selected variables were identified by literature review of the factors that impact the frequency and severity of viral illnesses, including COVID-19. The following variables were collected: age, gender, underinsured population, ethnicity, influenza vaccination status, population density (persons per square mile), urban air quality rank (lower number signifying better air quality), drinking water quality rank (lower number signifying better drinking water quality), ultraviolet index, precipitation (in inches), temperature (in degree Fahrenheit), census average household income (in US$), per capita spending on healthcare (in US$) and high school graduation rate. To capture comorbid conditions, the prevalence of obesity, prevalence of smoking, prevalence of cocaine abuse, prevalence of marijuana abuse, alcohol consumption (gallons per person per year), prevalence of asthma, prevalence of diabetes, prevalence of chronic obstructive pulmonary disease, prevalence of myocardial infarction, prevalence of coronary artery disease, prevalence of hypertension, prevalence of hyperlipidemia, and presence of inactivity were collected. To capture immunosuppressed states, the annual incidence of new cancer and HIV cases per 100,000 population were collected. Finally, the social distancing score by global positioning satellite data, public transportation volume, number of incarcerated inmates, number of nursing home residents, and tourism rank (lower number implies more tourists) were collected. Of these, ultraviolet index, temperature, and precipitation were averages for March and April of 2020, whereas the remainder of data were collected from the most recent iteration for each state. Much of the data was collected from government sources, such as the Centers for Disease Control and the complete list of sources is provided

The collected data represents state-level and not patient-level data. The endpoints were divided amongst the authors and collected by everyone. The data for each endpoint was then verified by another author who did not primarily collect the data. Finally, values in the top and bottom 10^th^ percentile were identified and verified by a third author.

### Statistical Analyses

As the data was collected for each state and intended for state-level analyses, the absolute number of COVID-19 cases, tests, and mortality were converted to a frequency using the state population. The frequencies for all the endpoints were calculated per 1,000,000 population. The case frequencies were then used as the dependent variables in a series of single-independent variable linear regressions to determine the univariate association between case frequency and the other variables previously defined and served as the univariate analyses. Next, a stepwise multivariate regression was conducted with p-value of 0.05 or less required for inclusion into the final model. Of the resulting models, the one with the highest R-squared value was selected as the final model. The same process was repeated for testing frequency and percent mortality as the dependent variable. Collinearity analyses were run with all multivariate regressions.

All statistical analyses were done using the user-coded, syntax-based interface of SPSS Version 23.0. A p-value of 0.05 or less was considered was considered statistically significant. The use of the word significant throughout the manuscript refers to “statistically significant” unless explicitly specified otherwise. All statistical analyses were done at the state-level with state-level data. Analyses were not conducted at a patient-level with patient-level data. The subjects here were the 50 states. The age, gender, and comorbidity prevalence are not based on patient-specific data but rather the state prevalence.

## Results

### COVID-19 Cases, Testing, and Mortality

A total of 1,090,500 COVID-19 cases were reported in the study period. This resulted in a case frequency of approximately 3,332 per 1,000,000 patients (3.3%). In the same period, 6,299,143 tests were done, resulting in a testing frequency 19,193 per 1,000,000 patients out of which 17.3% were reported positive. There were 63,642 deaths during this period which resulted in a mortality of 5.8%. Figures 1 and 2 demonstrate the case frequency and the percent mortality by state.

**Figure 1:**
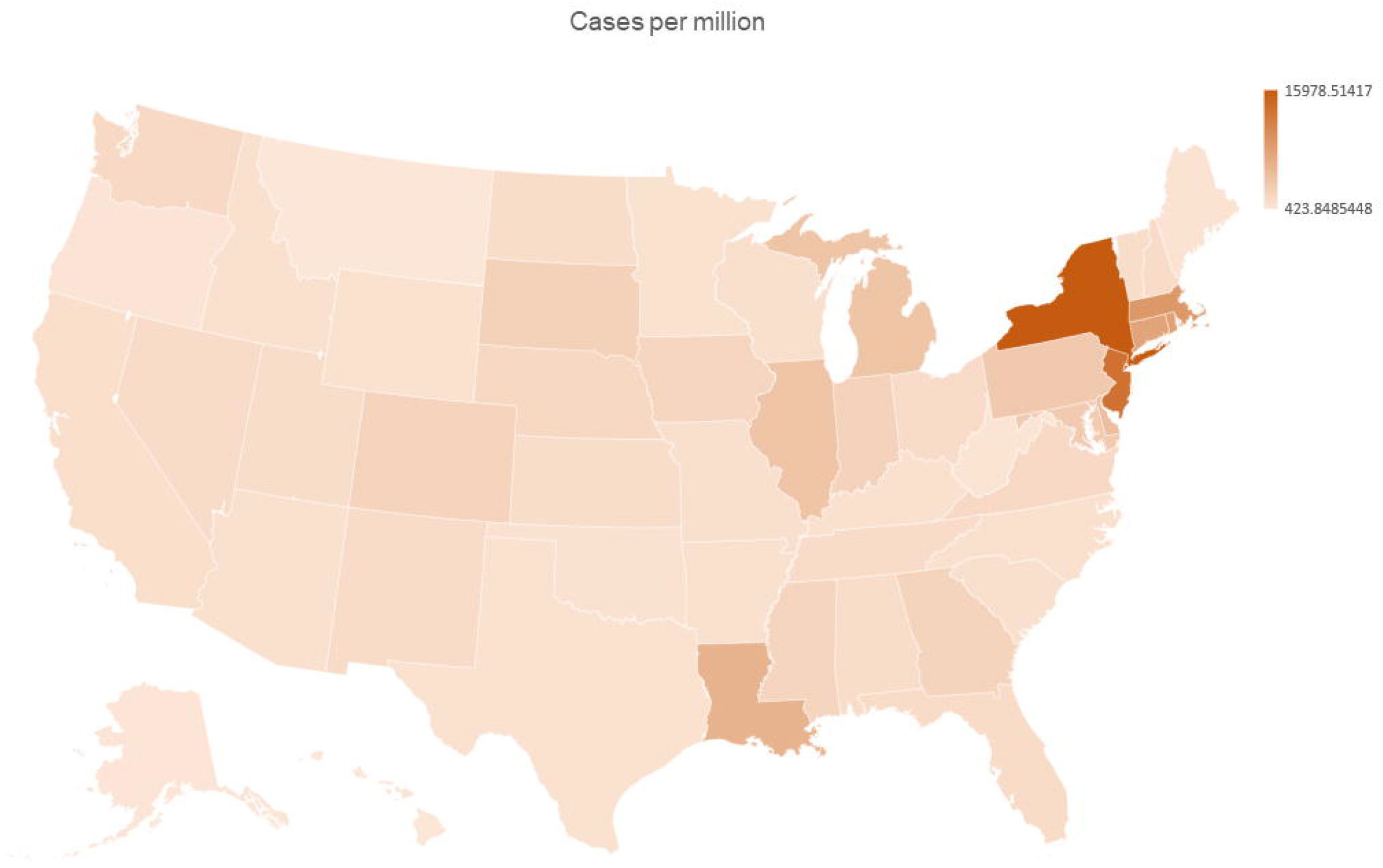
Cases per million in the United States divides by state.

**Figure 2:**
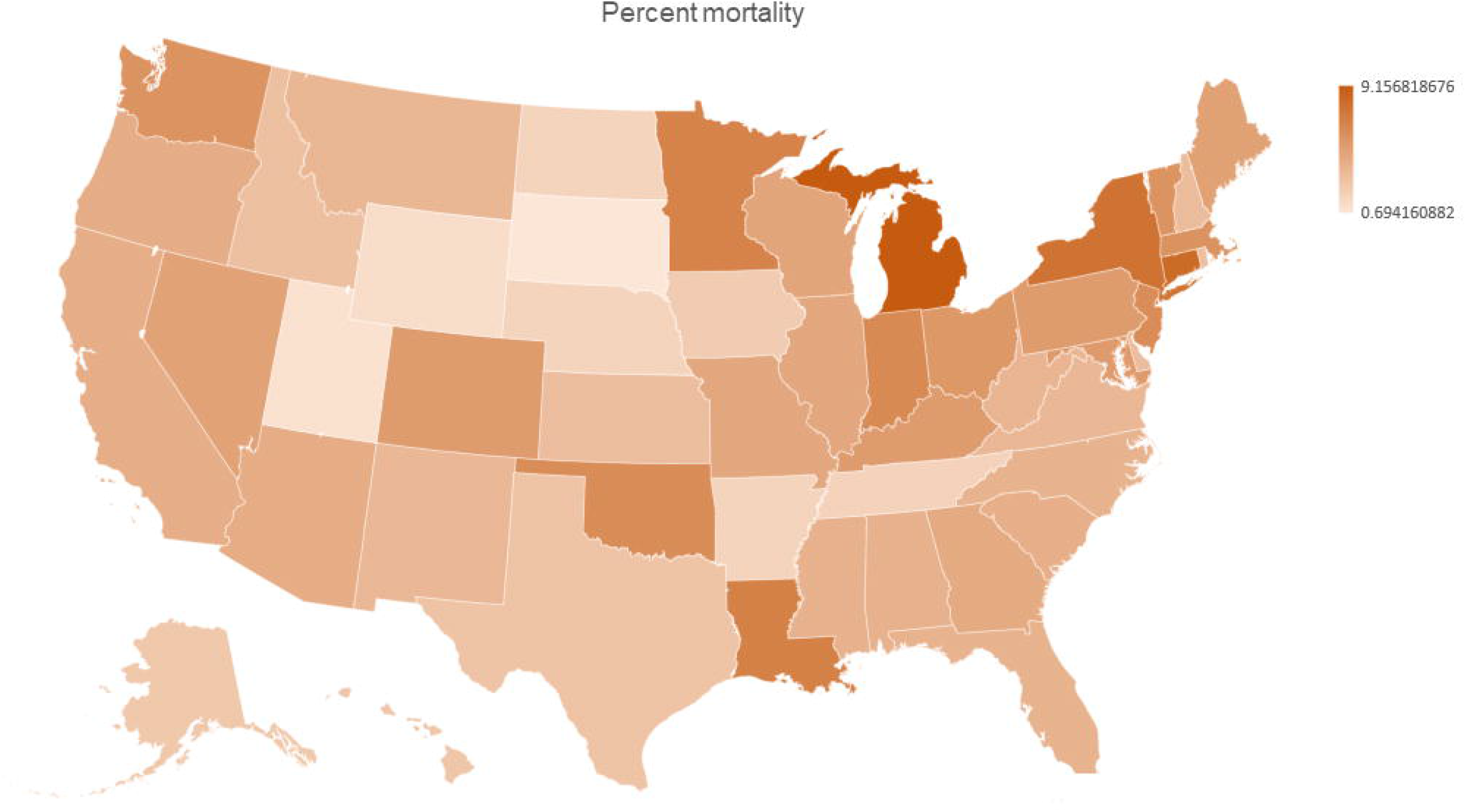
Percent mortality in the United States divided by state.

### COVID-19 Case Frequency, Univariate Analyses

The following factors were associated with greater case frequency on univariate linear regression analyses: female gender (beta-coefficient 1,095.8, p=0.03), higher population density (beta-coefficient 8.9, p< 0.01), lower ultraviolet index (beta coefficient -825.3, p=0.03), lower prevalence of obesity (beta coefficient -248.3, p=0.03), lower prevalence of uninsured (beta coefficient -298.6, p=0.01), higher frequency of other race (beta coefficient 38,638.7, p=0.01), lower prevalence of current smokers (beta coefficient -294.5, =0.02), higher per capita health care spending (beta coefficient 1.0, p< 0.01), higher public transportation volume (beta coefficient 0.1, p< 0.01), number of residents in nursing home facilities (beta coefficient 0.1, p< 0.01), and lower number for tourism ranking and thus more tourists (beta coefficient -79.8, p=0.01) (Table 1 shows full univariate data and Table 2 shows univariate results).

**Table 1:** Univariate analyses for factors associated with COVID-19 illness in the United States.

|  | **Cases per million** | **Tests per million** | **Mortality percent** |
| --- | --- | --- | --- |
| Age | 194.0 (p=0.31) | -61.7 (p=0.92) | 0.2 (p=0.01*) |
| Gender | 1095.8 (p=0.03) * | 1,065.2 (p=0.54) | 0.5 (p=0.05) |
| Race, White | -4,715.4 (p=0.18) | -8,401.8 (p=0.46) | -1.6 (p=0.41) |
| Race, Black | 7,312.7 (p=0.12) | -1,715.4 (p=0.91) | 4.3 (p=0.11) |
| Race, Native American | -23,515 (p=0.13) | 28,602.2 (p=0.57) | -16.6 (p=0.06) |
| Race, Asian | 6,237.8 (p=0.44) | 15,722.4 (p=0.55) | 1.0 (p=0.82) |
| Race, Islander | -30,422.1 (p=0.34) | 20,730.6 (p=0.84) | -20.5 (p=0.26) |
| Race, Other race | 38,638.7 (p=0.01) * | 87,436.9 (p=0.08) | 9.4 (p=0.30) |
| Race, multiple race | -13,133.5 (p=0.34) | 5,794.8 (p=0.89) | -4.6 (p=0.57) |
| Uninsured | -298.6 (p=0.01) * | -900.9 (p=0.02) * | -0.1 (p=0.02) * |
| Obesity | -248.3 (p=0.03) * | -577.5 (p=0.12) | -0.05 (p=0.39) |
| Asthma | 94.2 (p=0.80) | 1,845.9 (p=0.12) | 0.4 (p=0.04) * |
| Diabetes | -143.6 (p=0.55) | -394.9 (p=0.60) | 0.1 (p=0.69) |
| Chronic Obstructive Pulmonary Disease (COPD) | -254.7 (p=0.21) | -557.3 (p=0.40) | 0.1 (p=0.60) |
| Myocardial infarction | -586.2 (p=0.16) | -827.7 (p=0.54) | -0.1 (p=0.80) |
| Coronary Artery Disease | -517.2 (p=0.21) | -1,566.3 (p=0.23) | 0.1 (p=0.61) |
| Hypertension | -77.1 (p=0.48) | -223.8 (p=0.52) | 0.1 (p=0.82) |
| Hyperlipidemia | -1.9 (p=0.99) | -372.6 (p=0.49) | 0.1 (p=0.24) |
| Cancer | 23.1 (p=0.08) | 7.3 (p=0.86) | 0.1 (p=0.16) |
| Stroke | -846.5 (p=0.12) | -2,146.2 (p=0.22) | -0.1 (p=0.94) |
| Human Immunodeficiency Virus (HIV) | 114.5 (p=0.10) | 19.4 (p=0.93) | 0.1 (p=0.11) |
| Physical inactivity | -58.6 (p=0.73) | -10.2.1 (p=0.85) | -0.1 (p=0.80) |
| Received influenza vaccination | -56.7, (p=-0.51) | -397.0 (p=0.14) | 0.1 (p=0.70) |
| State population density | 8.9 (p< 0.01) * | 19.2 (p<0.01) * | 0.1 (p=0.02) * |
| Urban air quality rank (lower number is better quality) | 22.4 (p=0.49) | 5.1 (p=0.96) | 0.1 (p=0.97) |
| Drinking water quality rank (lower number is better quality) | -50.4 (p=0.12) | -50.8 (p=0.63) | -0.1 (p=0.57) |
| Ultraviolet (UV) index | -825.3 (p=0.03) * | -1,429.2 (p=0.26) | -0.3 (p=0.09) |
| Precipitation | 103.7 (p=0.70) | -199.6 (p=0.82) | 0.1 (p=0.46) |
| Average temperature | -12.3 (p=0.76) | -159.7 (p=0.23) | -0.1 (p=0.88) |
| Average household income | 0.1 (p< 0.01) * | 0.1 (p=0.08) | 0.1 (p=0.09) |
| High school grad percent | -82.5 (P=0.58) | -75.1 (p=0.87) | -0.1 (p=0.52) |
| Social distancing score | 442.3 (p=0.57) | 1,986.2 (p=0.43) | 0.1 (p=0.77) |
| Alcohol consumption | -189.2 (p=0.81) | 251.7 (p=0.92) | -0.1 (p=0.68) |
| Current cigarette smoker | -294.5 (p=0.02) * | -683.0 (p=0.11) | -0.1 (p=0.50) |
| Cocaine | 754.2 (p=0.32) | 2,852.1 (p=0.24) | 1.0 (p=0.01) * |
| Marijuana | -9.1 (p=0.92) | 262.9 (p=0.41) | 0.1 (p=0.06) |
| Per capita health care spending | 1.0 (p< 0.01) * | 3.68 (p<0.01) * | 0.1 (p=0.18) |
| Public transportation volume | 0.1 (p<0.01) * | 0.1 (p=0.01) * | 0.1 (p=0.02) * |
| Number of residents in nursing home facilities | 0.1 (p<0.01) * | -0.1 (p=0.88) | 0.1 (p=0.01) * |
| Number of inmates in prisons | 0.1 (p=0.98) | -0.1 (p=0.11) | 0.1 (p=0.37) |
| Tourism ranking 2018 (lower ranking means more tourists) | -79.8 (p=0.01) | -50.9 (p=0.65) | -0.1 (p=0.02) * |

\* Statistically significant (p-value <0.05)

**Table 2:** Associations between demographic factors, comorbidities, and environmental factors on case numbers, test numbers and percent mortality in univariate analysis.

|  | **Cases per million** | **Tests per million** | **Mortality percent** |
| --- | --- | --- | --- |
| Age | No | No | Yes (more mortality with increased age) |
| Gender | Yes (higher frequency with more females) | No | No |
| Race, White | No | No | No |
| Race, Black | No | No | No |
| Race, Native American | No | No | No |
| Race, Asian | No | No | No |
| Race, Islander | No | No | No |
| Race, Other race | Yes (higher frequency with other race) | No | No |
| Race, multiple race | No | No | No |
| Uninsured | Yes (higher frequency with less uninsured) | Yes (lower frequency with more uninsured) | Yes (lower mortality with more uninsured) |
| Obesity | Yes (higher frequency with lower obesity) | No | No |
| Asthma | No | No | Yes (higher mortality with more asthma) |
| Diabetes | No | No | No |
| Chronic Obstructive Pulmonary Disease (COPD) | No | No | No |
| Myocardial infarction | No | No | No |
| Coronary Artery Disease | No | No | No |
| Hypertension | No | No | No |
| Hyperlipidemia | No | No | No |
| Cancer | No | No | No |
| Stroke | No | No | No |
| Human Immunodeficiency Virus (HIV) | No | No | No |
| Physical inactivity | No | No | No |
| Received influenza vaccination | No | No | No |
| State population | Yes (higher frequency | Yes (higher frequency | Yes (more |

| density | with more density) | with more density) | mortality with greater density) |
| --- | --- | --- | --- |
| Urban air quality rank (lower number is better quality) | No | No | No |
| Drinking water quality rank (lower number is better quality) | No | No | No |
| Ultraviolet (UV) index | Yes (higher frequency with lower UV index) | No | No |
| Precipitation | No | No | No |
| Average temperature | No | No | No |
| Average household income | Yes (higher frequency with higher average income) | No | No |
| High school grad percent | No | No | No |
| Social distancing score | No | No | No |
| Alcohol consumption | No | No | No |
| Current cigarette smoker | Yes (higher frequency with lower smoking) | No | No |
| Cocaine | No | No | Yes |
| Marijuana | No | No | No |
| Per capita health care spending | Yes (higher frequency with higher spending) | Yes (higher frequency with higher spending) | No |
| Public transportation volume | Yes (higher frequency with higher volume) | Yes (higher frequency with higher volume) | Yes (higher mortality with higher volume) |
| Number of residents in nursing home facilities | Yes (higher frequency with higher number) | No | Yes (higher mortality with higher number) |
| Number of inmates in prisons | No | No | No |
| Tourism ranking 2018 (lower number means more tourists) | Yes (higher frequency with more tourists) | Yes (higher frequency with more tourists) | Yes (higher mortality with more tourists) |

**Table 3:**
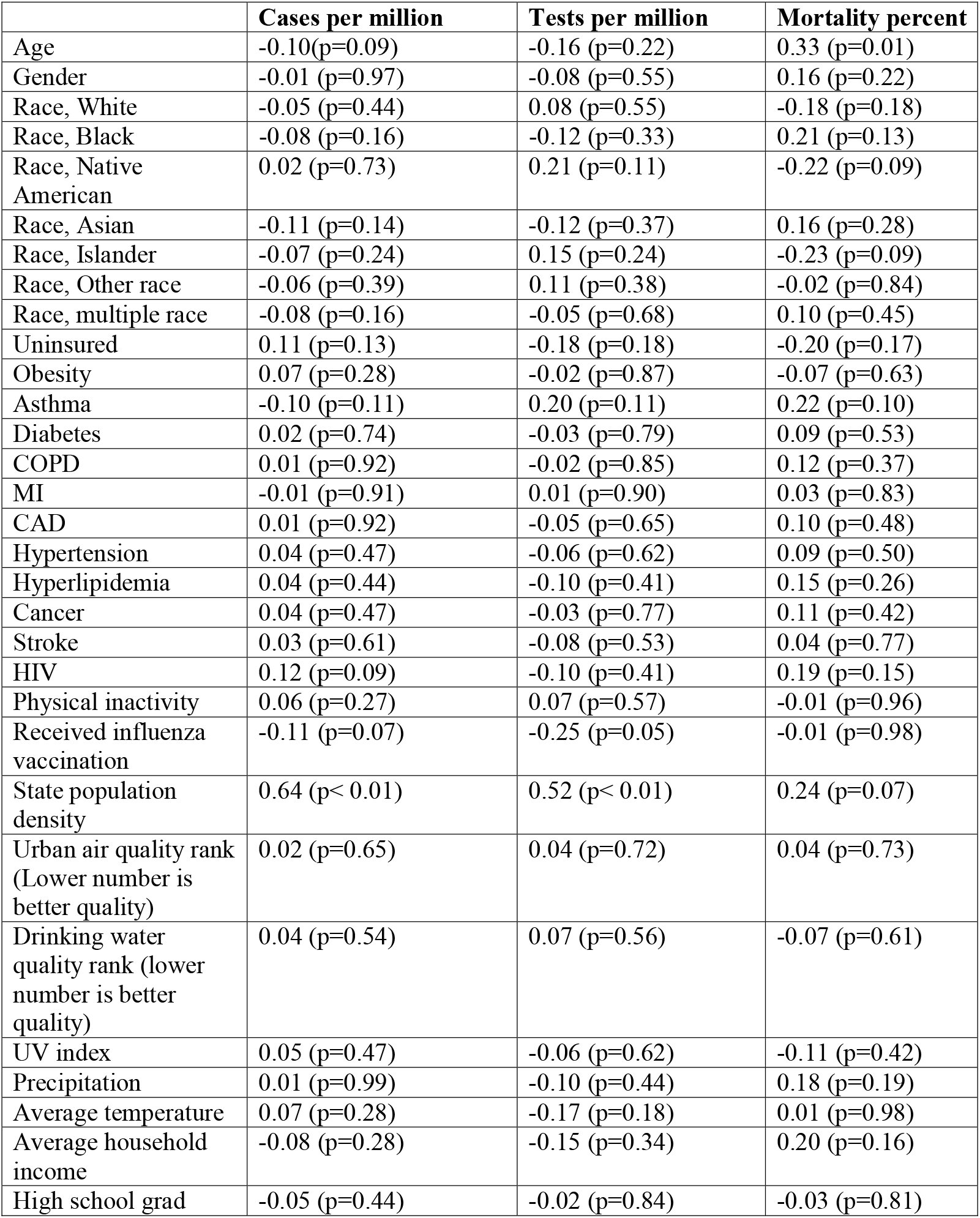
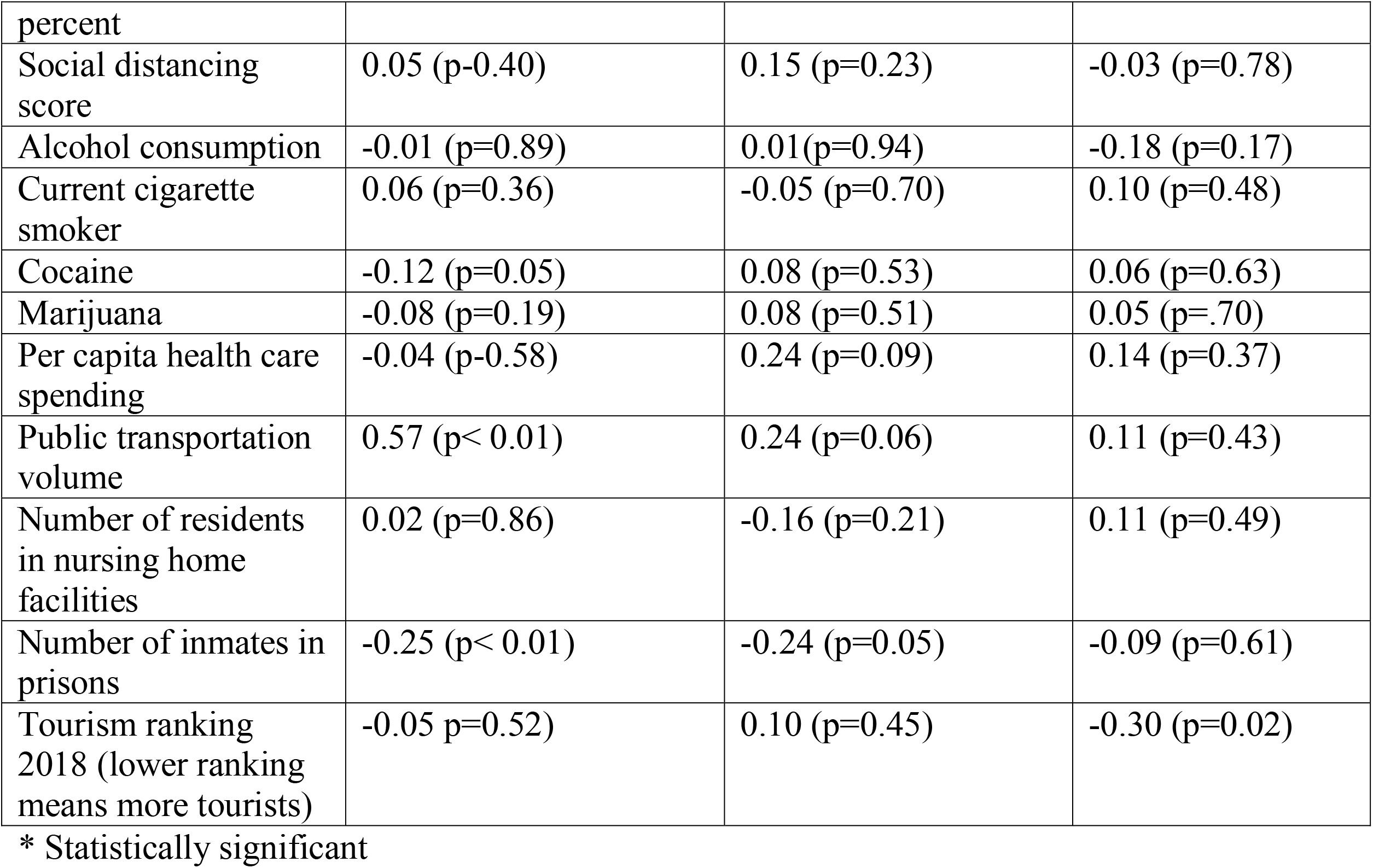
Associations between demographic factors, comorbidities, and environmental factors on case numbers, test numbers and percent mortality in multivariate analysis.

|  | **Cases per million** | **Tests per million** | **Mortality percent** |
| --- | --- | --- | --- |
| Age | -0.10(p=0.09) | -0.16 (p=0.22) | 0.33 (p=0.01) |
| Gender | -0.01 (p=0.97) | -0.08 (p=0.55) | 0.16 (p=0.22) |
| Race, White | -0.05 (p=0.44) | 0.08 (p=0.55) | -0.18 (p=0.18) |
| Race, Black | -0.08 (p=0.16) | -0.12 (p=0.33) | 0.21 (p=0.13) |
| Race, Native American | 0.02 (p=0.73) | 0.21 (p=0.11) | -0.22 (p=0.09) |
| Race, Asian | -0.11 (p=0.14) | -0.12 (p=0.37) | 0.16 (p=0.28) |
| Race, Islander | -0.07 (p=0.24) | 0.15 (p=0.24) | -0.23 (p=0.09) |
| Race, Other race | -0.06 (p=0.39) | 0.11 (p=0.38) | -0.02 (p=0.84) |
| Race, multiple race | -0.08 (p=0.16) | -0.05 (p=0.68) | 0.10 (p=0.45) |
| Uninsured | 0.11 (p=0.13) | -0.18 (p=0.18) | -0.20 (p=0.17) |
| Obesity | 0.07 (p=0.28) | -0.02 (p=0.87) | -0.07 (p=0.63) |
| Asthma | -0.10 (p=0.11) | 0.20 (p=0.11) | 0.22 (p=0.10) |
| Diabetes | 0.02 (p=0.74) | -0.03 (p=0.79) | 0.09 (p=0.53) |
| COPD | 0.01 (p=0.92) | -0.02 (p=0.85) | 0.12 (p=0.37) |
| MI | -0.01 (p=0.91) | 0.01 (p=0.90) | 0.03 (p=0.83) |
| CAD | 0.01 (p=0.92) | -0.05 (p=0.65) | 0.10 (p=0.48) |
| Hypertension | 0.04 (p=0.47) | -0.06 (p=0.62) | 0.09 (p=0.50) |
| Hyperlipidemia | 0.04 (p=0.44) | -0.10 (p=0.41) | 0.15 (p=0.26) |
| Cancer | 0.04 (p=0.47) | -0.03 (p=0.77) | 0.11 (p=0.42) |
| Stroke | 0.03 (p=0.61) | -0.08 (p=0.53) | 0.04 (p=0.77) |
| HIV | 0.12 (p=0.09) | -0.10 (p=0.41) | 0.19 (p=0.15) |
| Physical inactivity | 0.06 (p=0.27) | 0.07 (p=0.57) | -0.01 (p=0.96) |
| Received influenza vaccination | -0.11 (p=0.07) | -0.25 (p=0.05) | -0.01 (p=0.98) |
| State population density | 0.64 (p< 0.01) | 0.52 (p< 0.01) | 0.24 (p=0.07) |
| Urban air quality rank (Lower number is better quality) | 0.02 (p=0.65) | 0.04 (p=0.72) | 0.04 (p=0.73) |
| Drinking water quality rank (lower number is better quality) | 0.04 (p=0.54) | 0.07 (p=0.56) | -0.07 (p=0.61) |
| UV index | 0.05 (p=0.47) | -0.06 (p=0.62) | -0.11 (p=0.42) |
| Precipitation | 0.01 (p=0.99) | -0.10 (p=0.44) | 0.18 (p=0.19) |
| Average temperature | 0.07 (p=0.28) | -0.17 (p=0.18) | 0.01 (p=0.98) |
| Average household income | -0.08 (p=0.28) | -0.15 (p=0.34) | 0.20 (p=0.16) |
| High school grad | -0.05 (p=0.44) | -0.02 (p=0.84) | -0.03 (p=0.81) |
| percent |  |  |  |
| Social distancing score | 0.05 (p=0.40) | 0.15 (p=0.23) | -0.03 (p=0.78) |
| Alcohol consumption | -0.01 (p=0.89) | 0.01(p=0.94) | -0.18 (p=0.17) |
| Current cigarette smoker | 0.06 (p=0.36) | -0.05 (p=0.70) | 0.10 (p=0.48) |
| Cocaine | -0.12 (p=0.05) | 0.08 (p=0.53) | 0.06 (p=0.63) |
| Marijuana | -0.08 (p=0.19) | 0.08 (p=0.51) | 0.05 (p=.70) |
| Per capita health care spending | -0.04 (p=0.58) | 0.24 (p=0.09) | 0.14 (p=0.37) |
| Public transportation volume | 0.57 (p< 0.01) | 0.24 (p=0.06) | 0.11 (p=0.43) |
| Number of residents in nursing home facilities | 0.02 (p=0.86) | -0.16 (p=0.21) | 0.11 (p=0.49) |
| Number of inmates in prisons | -0.25 (p< 0.01) | -0.24 (p=0.05) | -0.09 (p=0.61) |
| Tourism ranking 2018 (lower ranking means more tourists) | -0.05 p=0.52) | 0.10 (p=0.45) | -0.30 (p=0.02) |

\* Statistically significant

### COVID-19 Case Frequency, Multivariate Analyses

The following factors were associated with greater case frequency on multivariable analyses: higher population density (beta coefficient 7.5, p< 0.01) and increased public transportation volume (beta coefficient 0.1, p< 0.01). The R-square for this model was 0.78. Collinearity analyses did not demonstrate any significant collinearity (Table 2 shows multivariate analyses).

### COVID-19 Testing Frequency, Univariate Analyses

The following factors were associated with greater testing frequency on univariate linear regression analyses: higher population density (beta coefficient 19.2, p< 0.01), lower prevalence of uninsured (beta coefficient -900.9, p=0.02), higher per capita health care spending (beta coefficient 3.68, p<0.01), and higher public transportation volume (beta coefficient 0.1, p=0.01).

### COVID-19 Testing Frequency, Multivariate Analyses

The following factors were associated greater testing frequency on multivariable analyses: higher population density (beta coefficient 19.9, p< 0.01). The R-square for this model was 0.27. Collinearity analyses did not demonstrate any significant collinearity.

### COVID-19 Percent Mortality, Univariate Analyses

The following factors were associated with greater percent mortality on univariate linear regression analyses: older age in years (beta coefficient 0.2, p=0.01), population density (beta coefficient 0.1, p=0.02), higher prevalence of asthma (beta coefficient 0.4, p=0.04), lower prevalence of uninsured (beta coefficient -0.1, p=0.02), higher prevalence of cocaine use (beta coefficient 1.0, p=0.01), higher public transportation volume (beta coefficient 0.1, p=0.02), higher number of residents in nursing home facilities (beta coefficient 0.1, p=0.01), and lower number for tourism ranking and thus more tourists (beta coefficient -0.1, p=0.02)

### COVID-19 Percent Mortality, Multivariate Analyses

The following factors were associated with greater percent mortality using multivariable analyses: median age in years (beta coefficient 0.2, p=0.01) and tourism ranking (beta coefficient-0.1, p=0.02). The R-square for this model was 0.29. Collinearity analyses did not demonstrate significant collinearity.

### Power Analyses

For multivariate regression analyses for case frequency, for which there is a relatively adequate effect size, and two predictors in the model, 31 subjects would be required to achieve 80% power. With 50 states and thus 50 subjects in these analyses, the multivariate analyses for case frequency are adequately powered.

For multivariable regression analyses for testing frequency, for which there is a relatively low effect size, and one predictor in the model, 385 subjects would be required to achieve 80% power. With 50 states and thus 50 subjects in these analyses, the multivariable analyses for testing frequency are not adequately powered.

For multivariable regression analyses for percent mortality, for which there is relatively low effect size, and two predictors in the model, 478 patients would be required to achieve 80% power. With 50 states and thus 50 subjects in these analyses, the multivariable analyses for percent mortality are not adequately powered.

## Discussion

In these analyses, wide variations in case frequency, testing frequency, and percent mortality were observed. These results also identified factors, such as population density, transportation volume, tourism index and older age to be some of the factors affecting the above outcomes. To the authors’ knowledge, this is the first study to incorporate large number of variables to study the differences in COVID-19 transmission among different states in the US.

Several laboratory and clinical risk factors have been postulated to predispose patients to symptomatic infection with COVID-19 and related mortality^7–9^. However, non-clinical factors that affect such outcomes are unknown. After multivariate analysis in this study, a higher population density was found to be associated with higher case and testing frequency. The results also identified increased public transportation to be associated with higher case frequency. Finally, the results identified older age and increased tourism independently related to higher mortality.

Since these analyses were based on state-level data, the power for multivariate regression was reduced compared to if the analyses were completed with patient-level data. Hence, the findings of the univariate analyses deserve attention as well here. We found direct relationships between case frequency in a state and female gender, underinsured status, average household income and per capita healthcare spending and inverse relationship with obesity, smoking and UV light exposure. Public transportation was significantly associated with frequency of cases, testing and mortality rates on univariate analysis. However, on multivariate analysis, only frequency of cases remained significantly associated with public transportation. Tourism ranking was also a significant predictor of all 3 endpoints on univariate analysis and remained significantly associated with mortality even after multivariate regression.

There is limited data on the nature of healthcare disparities during the COVID-19 pandemic. The cumulative COVID-19 incidence has been reported to be significantly variable among jurisdictions, ranging from 20.6 per 100,000 cases in Minnesota to 915.3 per 100,000 cases in New York City^10^. The timing of introduction of COVID-19 in the state and the extent of mitigation measures may mediate some of this variation. The age of patients has been shown to be a significant predictor of COVID-19 infection and worse outcomes^9,11^. The race and gender of patients have also been reported to be associated with a higher case frequency and worse outcomes in patients with COVID-19^12,13^. It continues to be shown that population density is associated with an increase in transmission and infection for the high risk population^14,15^.

The results of this study showed that the public transport volume was also linked to a higher case rate. Prior studies have demonstrated similar findings with influenza-like illnesses and that its use increases the individual’s risk for acquiring an acute respiratory infection^16,17^. A simulation model indicated that the high level of subway usage in New York can influence disease spread in an influenza epidemic and that between 4-5% of total infections would occur on subways^18^. This information is particularly important as recommendations to maintaining strict disinfecting guidelines for public transport along with shelter-in-place whenever possible are established^19,20^.

The results of this study identified that higher tourism is associated with increased mortality. A possible explanation of this identified phenomenon has been published and thought to be related to an influx of infected patients presenting late in the disease course^21,22^. Furthermore, prior studies have shown that air transportation accelerates viral spreading mainly related to high passenger traffic and risk of surface contamination in airports^23,24^. Similar findings have been reported in trains and other types of commercial vessels ^25,26^. Hence, the widespread implementation of travel restrictions, social distancing and lockdowns have become the main preventative interventions to decrease viral spreading during this pandemic. However, in this analysis, social distancing score was not associated with COVID-19 case frequency or mortality. The result from prior studies showed that social distancing is an effective measure at decreasing viral spreading when comprised of quarantines, school closure and workplace distancing^27^,^28^. However, our current analysis showed a lack of association between social distancing and COVID-19 case frequency, which may also represent a limitation of the process of measuring the social distancing score. This is particularly important as some countries like Sweden have encountered higher COVID-19 case frequencies after adopting more lenient social distancing measures^29^. This analysis also showed a lack of impact of climate-related factors on case and testing frequency. These findings require further validation as conflicting reports have been published^5,30,31^. Surprisingly, in our analysis states with higher number of uninsured patients were found to have lower mortality which could possibly be related to underreporting in such populations as both case numbers and testing numbers were also lower in such states.

Finally, most of the comorbidities analyzed were not found to be independently associated with the case mortality in this analysis. This may suggest the complex interplay between demographics, environmental factors and diseases processes^32,33^. The findings from this analysis also highlight some of the potential limitations of state-level data rather than patient-level data, as previous studies have found a few comorbidities to be associated with increases in case frequency and percent mortality.

These analyses offer some early assessment of the factors that may be mediating COVID-19 case frequency, testing frequency, and percent mortality. These analyses present associations using state-level data and not patient-level data. While these analyses offer novel data regarding case frequency, testing frequency, and percent mortality in the US, these analyses are not without their limitations. First, all the study data was captured from publicly available sources which only had data until 2018. The use of state-level data reduced the power of analyses, as we used for the multivariate regression models the number of states as the subjects. Although the data collection carries a risk of bias, this was minimized by utilizing multiple investigators for accuracy of data captured. Due to data unavailability other important outcomes such as case positivity rate could not be assessed. The ecologic design of the paper and use of various data sources with varying methods are other limitations of our study. As the pandemic continues some of the variables selected for this study may be subject to change, particularly as state resources may become overwhelmed. Finally, lack of granularity to county or city level data further limits our interpretations.

With these limitations in mind, it is important to frame the intentions of this study appropriately. These analyses are by no means intended to be definitive data but are intended to be exploratory data to help identify variables that should be accounted for in larger, multicenter studies that utilize patient-level data. Factors such as the environmental and local infrastructural characteristics appear to modulate the case frequency and percent mortality and thus could be beneficial to capture in future studies. The data from those variables may assist with the understanding of viral spreading and the pandemic evolution. For instance, the identification of the association between higher tourism volume with higher case frequency and percent mortality may help implement faster travel restrictions for future pandemics. Similarly, the association between public transportation volume and its association with increased case frequency and percent mortality may assist in developing future public response.

## Conclusion

This observational analysis of publicly reported state-level data identified factors associated with increased case frequency, testing frequency, and percent mortality for COVID-19. These data can guide future study design and develop risk prediction models.

## Data Availability

Deidentified individual participant data will be made available at https://figshare.com/articles/dataset/Case_Numbers_and_Mortality_in_US_Data_Extraction_xlsx/12730919

## Acknowledgement

None

## In Memoriam

We dedicate this paper to the memory of Luis Carlos Gamboa Chavez (1974-2020), devoted father, son, and friend.

## Figure and table legends

**Supplementary File 1**. Data sources

